# Stereo-electroencephalography Performance in Bilateral Independent/Unclear Scalp Seizures

**DOI:** 10.1101/2025.06.17.25329726

**Authors:** Aayesha J Soni, M Claudia Burbano, R Grace Couper, Poul H Espino, John AL Perez, Amit Persad, Khalid Alorabi, Poornima N Nambiar, Arun Thurairajah, David Diosy, Michelle-Lee Jones, Ana Suller Marti, Keith W MacDougall, Jonathan C Lau, David A Steven, Jorge G Burneo, Giovanni Pellegrino

## Abstract

Scalp electroencephalography (EEG) may reveal bilateral independent or unclear (BI/U) ictal onset patterns in patients with focal drug-resistant epilepsy, presenting a challenge to surgical decision-making. The utility of stereo-electroencephalography (SEEG) in this subgroup, particularly the probability of delineating a single seizure onset zone (SOZ) that would permit curative resection, remains poorly understood. This study examined whether BI/U scalp EEG findings could predict SEEG outcomes in this population.

We conducted a retrospective cohort study of consecutive patients with focal drug-resistant epilepsy and BI/U ictal onset on scalp EEG who underwent SEEG evaluation at the London Health Sciences Centre (Ontario, Canada) between January 2012 and December 2024. All patients had undergone non-invasive and invasive presurgical assessments. Surgical outcomes were determined using the Engel classification following at least one year of postoperative follow-up. A blinded decision validation sub-study was also performed. Blinded to actual outcomes, the team made decisions regarding SEEG and surgical interventions when patients found to have a single SEEG SOZ were presented. Responses were stratified to inform the added diagnostic value of SEEG.

Of 255 SEEG cases screened, 84 patients (33%) met inclusion criteria. The cohort was 56% female, with a median seizure onset age of 12 years (IQR 6–20); 65.5% had temporal lobe epilepsy (TLE). A single SOZ was identified in 14.3% of cases (TLE: 14.5%, extratemporal: 13.8%). Patients with a single SEEG SOZ were found to have shorter recording durations (mean of 11 vs 15.79 days in those with multifocal SEEG SOZs; p=0.009). Curative focal resections were performed in 12% (*n=*10), with long-term Engel I outcomes achieved in one patient (1.2%). Palliative resections occurred in 26% (*n=*22), with Engel I outcomes in 7% (*n=*6). In 50% of the blinded cases, the epilepsy surgery team reported that they would not have recommended SEEG based on phase I data.

These findings suggest that patients with BI/U scalp EEG SOZs may be associated with a low likelihood of identifying a single SEEG SOZ and curative outcome. Using BI/U scalp EEG ictal onset as a predictor in preoperative decision-making will assist in refining SEEG candidate selection in this large subgroup.

## Introduction

In selected patients with focal drug resistant epilepsy, resective surgery is an effective and safe option to achieve seizure control and improve quality of life. ^1–3^ This is conditional on identifying a unilateral, single, and spatially restricted region from where seizures arise, identified as the seizure onset zone (SOZ). To this end, non-invasive presurgical planning involves the assimilation of information obtained from the epilepsy history, semiology, scalp/video electroencephalography (EEG) monitoring, brain magnetic resonance imaging (MRI), ^4^ neuropsychological evaluation, and various other ancillary investigations. In certain complex cases, these phase I investigations result in one or more epilepsy localization hypotheses that require stereo-electroencephalography (SEEG) to better delineate the potential surgical target. SEEG may provide crucial information to offer beneficial resective surgery and has been increasingly utilized. ^5^

During the phase I investigation of a potential epilepsy surgery candidate, scalp EEG may demonstrate an ictal onset arising independently from both hemispheres or of an unclear onset. Cases of these patients having a unilateral focus have been documented, warranting consideration of a phase II SEEG investigation. Examples include a midline generator, a small focus, a deep generator or rapid propagation to both hemispheres from a single generator. ^6, 7^ SEEG is pursued in these cases under the hypothesis that seizures are only seen on scalp EEG when there is sufficient extent and synchrony of cortical activation, ^8^ and that if these criteria are unmet by the limited spatial sampling of scalp EEG recordings then seizures may present with a bilateral independent/unclear (BI/U) onset. Outcomes in this group have been shown to be worse following resective surgery, as opposed to SEEG patients with preimplantation hypotheses restricted to one hemisphere. ^9^ Furthermore, up to a quarter of patients undergoing SEEG are not candidates for resective neurosurgery. ^10, 11^ Of those who are, up to a half may not maintain seizure freedom. ^12^ Hence, identifying appropriate candidates for SEEG is nuanced; though SEEG and epilepsy surgery should be offered to maximize quality of life, undue surgical morbidity and healthcare costs should be minimized.

Retrospective reports describe general outcomes following SEEG implantation, ^13, 14^ but there is no data on the yield of identifying a single, unilateral SOZ following SEEG implantation in patients with a BI/U scalp EEG ictal onset. Assessing patients for epilepsy surgery is an individualized decision comprising information from many tests. Acknowledging this, our aim is to evaluate the use of BI/U scalp EEG SOZs as a single predictor in determining the value of an SEEG implantation in this subgroup. The results will better inform pre-operative patient selection and counselling.

## Methods

### Study Design, Setting and Variables

This study evaluated the data in two separate ways. First, we analyzed our single centre retrospective cohort, and then a blinded decision validation sub-study using a simulated epilepsy surgery conference was conducted.

#### Observational Cohort Study

The observational cohort analysis was conducted using the Epilepsy Surgery Database at our institution. Adult patients (aged >17 years) with focal drug-resistant epilepsy who underwent an epilepsy surgery evaluation at the London Health Sciences Centre between January 2012 and December 2024 were considered for inclusion. From this cohort, individuals who had undergone SEEG were identified. Their epilepsy surgery conference documentation was retrospectively reviewed to determine eligibility. Inclusion was limited to patients whose scalp EEG demonstrated BI/U ictal onset patterns. These were defined as seizures which began either independently in both hemispheres or with non-localizable (normal), diffuse, or multifocal onsets that could not be confidently lateralized. Classification of BI/U ictal onset required documented consensus during the epilepsy surgery conference of these electrographic patterns as the predominant scalp EEG SOZ during clinical and/or electrographic seizures. Included in the BI group were patients with evidence of independent bitemporal SOZs. Seizure outcomes were obtained from the most recent visit in the epilepsy clinic, stratified according to the Engel classification system. ^15^ Only patients with a follow-up of longer than one year were included in this outcome analysis.

Demographic data and preoperative clinical information, sex, handedness, age at epilepsy diagnosis, seizure types, etiology of epilepsy, predominant scalp EEG ictal onset, neuropsychology and MRI brain results (type and location of lesion) were collected, along with ancillary information (FDG-PET, CT brain SPECT) whenever available. Phase II variables included age at SEEG insertion, number of SEEG electrodes inserted and duration of monitoring, and SEEG ictal, interictal and cortical stimulation findings. Interventions included the details of focal resective surgery, neuromodulation or radio-frequency thermo-coagulation (RF-TC) with the type, localization and intent. The duration of follow up and Engel outcome at the last visit was also documented. (Supplementary material)

#### Blinded Decision Validation Sub-study

A blinded decision-validation sub-study was also undertaken to assess the consistency of clinical decision-making and to evaluate the added diagnostic value of SEEG in this complex subgroup. Clinically relevant information from the phase I and II evaluations of patients who had a single SEEG SOZ was collected. An epilepsy surgery conference like the routine conferences was held, with two epileptologists and two functional neurosurgeons. They were presented with the phase I evaluation and asked to make one of the following decisions: stop (no further surgery offered), implantation of SEEG, operation with curative resective surgery or palliative surgery/neuromodulation. They were then provided with the information regarding the phase II evaluation, including the SEEG electrode coverage, ictal and inter-ictal findings. Again, they were asked to make an interventional decision. After being provided with the details regarding what intervention the patient had as well as the outcome, they were finally asked whether they would have changed any of their management decisions, and why/why not. Decisions for both phases of evaluation were recorded, which allowed us to define the proportion of patients in whom the management plan was changed and assess the additional diagnostic value of SEEG in these cases.

The reporting of this study followed Strengthening the Reporting of Observational studies in Epidemiology (STROBE) guidelines. ^16^ The study protocol was approved by the Western University Health Science Research Ethics Board (127171-108606) in London, Ontario, Canada.

### Outcome Measures

The primary outcome was the likelihood of a patient with BI/U scalp EEG SOZs having a single SOZ on SEEG. Successful identification of a single focus is defined as unambiguous SEEG electroclinical onset within a defined sub-lobar area, as documented by multidisciplinary conference consensus, regardless of any subsequent surgical treatment. Secondary outcome measures included the probability of subsequent focal resective surgery and tendency of seizure freedom as a function of preoperative and SEEG intervention factors. In addition, assessing the congruence and additional diagnostic value of an epilepsy surgery team identifying which patients should proceed to an SEEG implantation were measured. We differentiated between the intent of focal resective surgery as either curative or palliative, as documented in the notes of the epilepsy surgery conference of these patients. When performed with curative intent, the surgery was performed with the aim to achieve long-term seizure freedom by excising a discrete, well-localized, and functionally non-eloquent epileptogenic region. Conversely, palliative focal resective surgery was undertaken when sustained seizure freedom was unlikely and the primary surgical goal was improving seizure burden, reducing seizure severity, or enhancing quality of life.

### Statistical Methods

Demographic variables were described using means and standard deviations for normally distributed continuous variables, and medians with interquartile ranges for non-normally distributed continuous variables. Frequencies and percentages were used for categorical variables. To compare frequencies across groups, Chi-squared tests or Fisher’s Exact tests were used for categorical variables, and t-tests or Mann-Whitney U tests were used for continuous variables.

### Data Availability

The data supporting the findings of this study are available from the corresponding author upon reasonable request.

## Results

### Cohort Description and Phase I Investigation

A total of 84/255 (33%) consecutive patients who had undergone an SEEG investigation over 13 years met the inclusion criteria. (Figure 1)

**Figure 1.**
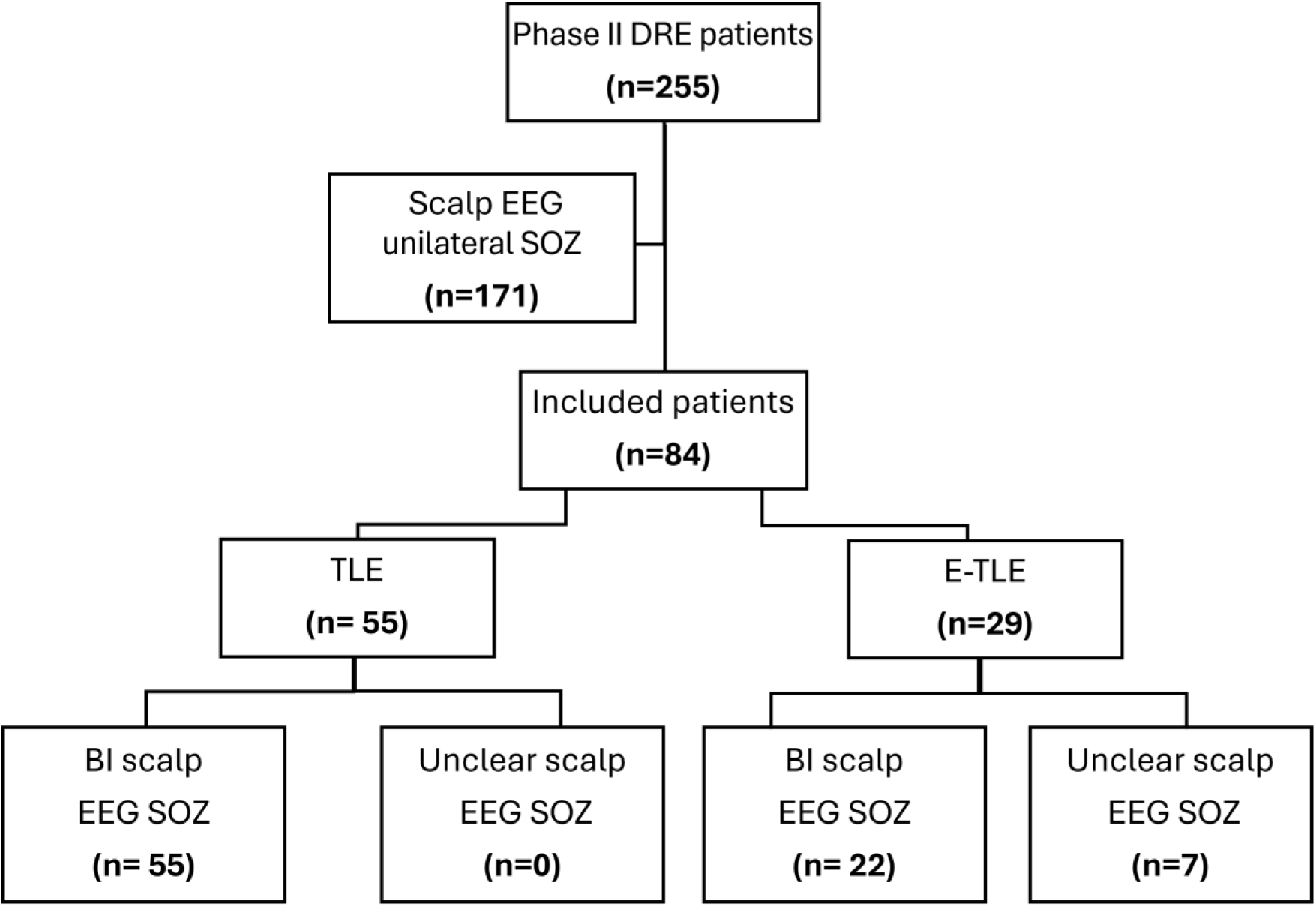
Patient Selection Flow Chart. The SEEG database over 13 years was assessed, and relevant patients included. They were then further characterized into TLE and E-TLE based on phase I data, allowing comparison between the two groups. DRE= drug resistant epilepsy; EEG= electroencephalogram; BI/U=bilateral independent/unclear; SOZ= seizure onset zone; TLE= temporal lobe epilepsy; E-TLE= extratemporal lobe epilepsy.

Table 1 documents details of all demographic and phase I non-invasive data collected, comparing patients who were initially classified as having temporal lobe epilepsy (TLE) versus extratemporal lobe epilepsy (E-TLE).

**Table 1.**
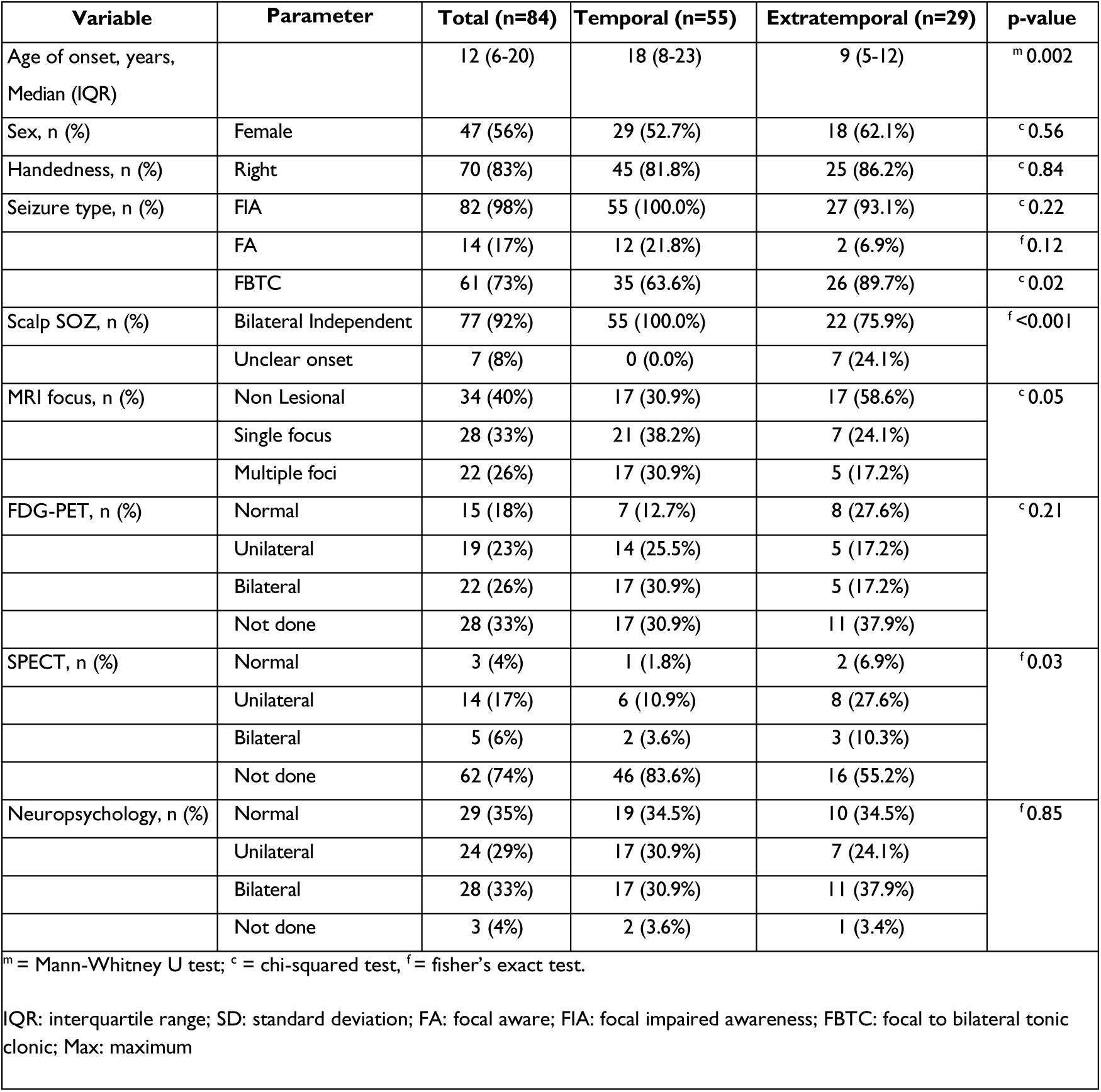
Details of demographic and phase I non-invasive investigations of all patients. Results have been divided into temporal and extra-temporal lobe epilepsy.

### Phase II SEEG Investigation

Table 2 documents the phase II investigation results together with intervention factors, divided again into the TLE and E-TLE groups for comparison. All patients were initially implanted with SEEG for a diagnostic purpose, as opposed to a therapeutic goal (e.g. RF-TC). Figure 2 is an illustrative case from our cohort.

**Table 2.** Details of phase II Invasive investigations as well as subsequent interventions and outcomes. Results have been divided into temporal and extra-temporal lobe epilepsy subgroups.

| Variable | Parameter | Total (n=84) | Temporal (n=55) | Extratemporal (n=29) | p-value |
| --- | --- | --- | --- | --- | --- |
| Age of SEEG, years, Median (IQR) |  | 32 (24-42) | 34 (26.5-45.5) | 27 (22-32) | <sup>m</sup>0.001 |
| SEEG SOZ, n (%) | Single focus | 12 (14%) | 8 (14.5%) | 4 (13.8%) | <sup>f</sup>1.00 |
|  | Multiple foci | 69 (82%) | 44 (80.0%) | 25 (86.2%) |  |
|  | Unknown | 3 (4%) | 3 (5.5%) | (0.0%) |  |
| SEEG interictal activity, n (%) | Max at seizure onset | 16 (19%) | 10 (18.2%) | 6 (20.7%) | <sup>f</sup>0.78 |
|  | Multiple foci | 68 (81%) | 45 (81.8%) | 23 (79.3%) |  |
|  | Single focus | 0 (0%) | 0 (0%) | 0 (0%) |  |
| SEEG cortical stimulation performed, n (%) |  | 51 (61%) | 31 (56.4%) | 20 (69.0%) | <sup>c</sup>0.37 |
| SEEG Stimulation-induced seizures, n (%) | Unilateral | 18 (21%) | 9 (16.4%) | 9 (31.0%) | <sup>c</sup>0.29 |
|  | Bilateral | 20 (24%) | 15 (27.3%) | 5 (17.2%) |  |
|  | No Seizures | 14 (17%) | 8 (14.5%) | 6 (20.7%) |  |
|  | Not done | 32 (38%) | 23 (41.8%) | 9 (31.0%) |  |
| RFTC, n (%) |  | 10 (12%) | 5 (9.1%) | 5 (17.2%) | <sup>c</sup>0.46 |
| Resective curative surgery, n (%) |  | 10 (11.9%) | 6 (10.9%) | 4 (13.8%) | <sup>f</sup>0.73 |
| Post-surgery follow-up >1 year, n (%) * |  | 6 (60%) | 2 (33.3%) | 4 (100.0%) | <sup>f</sup>0.08 |
| Engel classification, n (%) * | Class I | 1 (17%) | 1 (50.0%) | 0 (0.0%) | <sup>f</sup>1.00 |
|  | Class II | 1 (17%) | 0 (0.0%) | 1 (25.0%) |  |
|  | Class III | 1 (17%) | 0 (0.0%) | 1 (25.0%) |  |
|  | Class IV | 3 (50%) | 1 (50.0%) | 2 (50.0%) |  |
| Palliative surgery, n (%) |  | 22 (26%) | 12 (21.8%) | 10 (34.5%) | <sup>c</sup>0.32 |
| Post-surgery follow-up >1 year, n (%) * |  | 17 (77%) | 9 (75.0%) | 8 (80.0%) | <sup>c</sup>0.82 |
| Engel classification, n (%) * | Class I | 6 (35%) | 3 (33.3%) | 3 (37.5%) | <sup>f</sup>0.50 |
|  | Class II | 2 (12%) | 2 (22.2%) | 0 (0.0%) |  |
|  | Class III | 4 (24%) | 2 (22.2%) | 2 (25%) |  |
|  | Class IV | 5 (29%) | 2 (22.2%) | 3 (37.5%) |  |
| Neuromodulation, n (%) | Total | 41 (49%) | 28 (50.9%) | 13 (44.8%) | <sup>c</sup>0.76 |
| Neuromodulation type, n (%) * | VNS | 36 (88%) | 25 (89.3%) | 11 (84.6%) | <sup>f</sup>0.08 |
|  | RNS | 3 (7%) | 3 (10.7%) | 0 (0.0%) |  |
|  | DBS | 2 (5%) | 0 (0.0%) | 2 (15.4%) |  |

<sup>m</sup> = Mann-Whitney U test; <sup>c</sup> = chi-squared test, <sup>f</sup> = fisher's exact test, \* denominators of rows above

**Figure 2.**
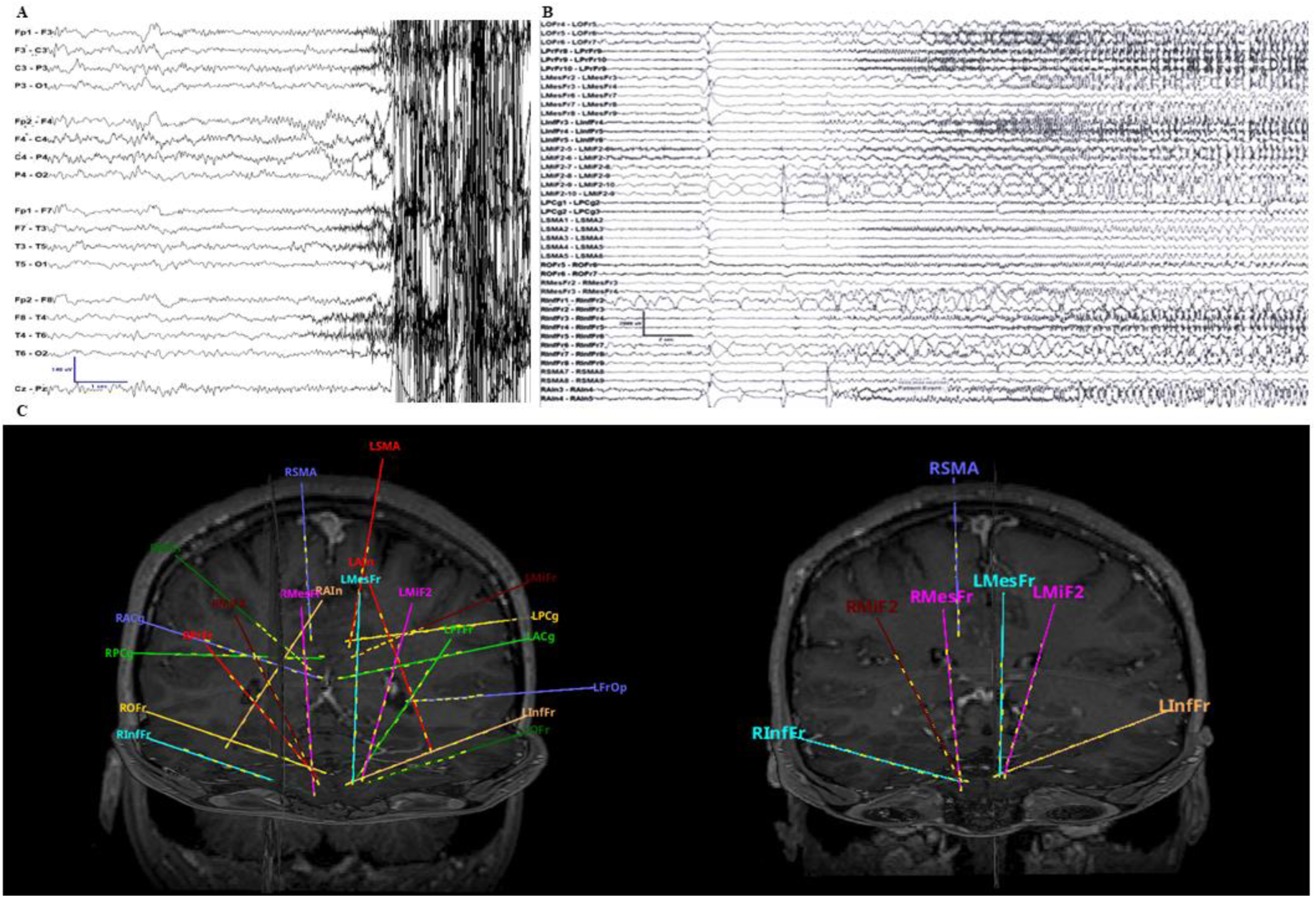
Illustrative case: scalp EEG and SEEG seizure onset, and SEEG implantation. (A) Unclear scalp EEG seizure onset, followed by diffuse muscle artifact that obscures the recording. (B) SEEG recording reveals seizure onset as a broad baseline shift, with subsequent diffuse attenuation, involving bihemispheric electrodes. (C) Post-implant SEEG visualization: 3D rendering of electrode placement. *Left:* all implanted electrodes (n = 22); *Right:* the additional six electrodes added to further localize the seizure onset zone. Despite this, the seizure onset could not be clearly delineated. EEG = Electroencephalography; SEEG = stereo-electroencephalography.

A single SEEG SOZ was identified in 12/84 patients (14.3%), with comparable rates in TLE (8/55; 14.55%) and E-TLE (4/29; 13.7%; p > 0.200). Of note, all TLE patients had a BI scalp ictal onset pattern. Out of the four E-TLE patients found to have a single SEEG SOZ, it was proposed that one had a deep focus and three had a mesial generator. The case with a deep focus originated from the insula, while the three midline generators included the precuneus, posterior supplementary sensorimotor area and mesial orbitofrontal lobes. Among E-TLE patients with bilateral independent scalp EEG SOZs, only 1/22 (4.55%) had a single focus, compared to 3/7 (42.86%) of those with unclear EEG onset (Fischer’s exact test = 6.555, p = 0.034). In all three cases, the unclear ictal onset on scalp EEG met our predefined criteria for non-localizable patterns and corresponded with patients in whom a mesial focus was proposed after the SEEG investigation.

Table 3 documents the differences in pre-surgical variables between patients with a single SEEG SOZ and those with multifocal/unknown SOZs. Variables with missing values were excluded. The duration of SEEG was also significantly different between those with single and multifocal/unknown SEEG SOZs, when all patients were included and when patients who pulled out electrodes before their monitoring could be completed were excluded. Patients with a single SEEG SOZ tended to have shorter SEEG monitoring durations. Additionally, patients with a single SEEG SOZ had more SEEG interictal activity present at the SOZ.

**Table 3.** Details of the differences in pre-surgical variables between patients with a single SEEG SOZ and those with multifocal/unknown SOZ. Variables with missing values were excluded.

| Variable | Parameter | Single SEEG SOZ (n=12) | Multi/unknown SEEG SOZ (n=72) | p-value |
| --- | --- | --- | --- | --- |
| Age at onset (median, IQR) |  | 12 (9.75-18.25) | 12 (5.75-20.25) | <sup>m</sup> 0.96 |
| Sex | Female | 7 (58.3%) | 40 (55.6%) | <sup>c</sup> 1.00 |
| Handedness | Right | 12 (100%) | 58 (80.6%) | <sup>c</sup> 0.21 |
| Seizure type | FIA | 12 (100%) | 70 (97.2%) | <sup>c</sup> 1.00 |
|  | FA | 0 (0%) | 14 (19.4%) | <sup>f</sup> 0.20 |
|  | FBTC | 11 (91.7%) | 50 (69.4%) | <sup>c</sup> 0.21 |
| Scalp EEG Onset | Bitemporal | 9 (75.0%) | 68 (94.4%) | <sup>f</sup> 0.06 |
|  | Unclear | 3 (25.0%) | 4 (5.6%) |  |
| MRI | Negative | 4 (33.3%) | 30 (41.7%) | <sup>f</sup> 0.12 |
|  | Single | 7 (58.3%) | 21 (29.2%) |  |
|  | Multifocal | 1 (8.3%) | 21 (29.2%) |  |
| FDG-PET, n (%) | Normal | 1 (8.3%) | 14 (19.4%) | <sup>f</sup> 0.72 |
|  | Unilateral | 4 (33.3%) | 15 (20.8%) |  |
|  | Bilateral | 3 (25.0%) | 19 (26.4%) |  |
|  | Not done | 4 (33.3%) | 24 (33.3%) |  |
| SPECT, n (%) | Normal | 1 (8.3%) | 2 (2.8%) | <sup>f</sup> 0.61 |
|  | Unilateral | 1 (8.3%) | 12 (16.7%) |  |
|  | Bilateral | 1 (8.3%) | 5 (6.9%) |  |
|  | Not done | 9 (75.0%) | 53 (73.6%) |  |
| Neuropsychology, n (%) | Normal | 5 (41.7%) | 24 (33.3%) | <sup>f</sup> 0.29 |
|  | Unilateral | 1 (8.3%) | 21 (29.2%) |  |
|  | Bilateral | 5 (41.7%) | 25 (34.7%) |  |
|  | Not done | 1 (8.3%) | 2 (2.8%) |  |
| Age at SEEG (median, IQR) |  | 28.5 (23-32.5) | 32 (24-44) | <sup>m</sup> 0.11 |
| Number of electrodes (mean, SD) |  | 12.33 (3.80) | 13.56 (3.11) | <sup>t</sup> 0.31 |
| Duration of electrodes (days) (mean, SD) (all patients) |  | 11 (4.99) | 15.79 (6.62) | <sup>t</sup> 0.009 |
| Duration of electrodes (days) (mean, SD) (limited patients) |  | 11.73 (4.52) | 16.14 (6.38) | <sup>t</sup> 0.01 |
| SEEG Interictal | Multifocal | 3 (25.0%) | 65 (90.3%) | <sup>f</sup> <0.001 |
|  | Max at SOZ | 9 (75.0%) | 7 (9.7%) |  |
| SEEG Stimulation |  | 8 (33.3%) | 43 (59.7%) | <sup>c</sup> 0.89 |
| SEEG Seizures | Not done | 4 (33.3%) | 28 (38.9%) | <sup>f</sup> 0.78 |
|  | Unilateral | 5 (41.7%) | 15 (20.8%) |  |
|  | Bilateral | 2 (16.7%) | 18 (25.0%) |  |
|  | None | 3 (25.0%) | 11 (15.3%) |  |
| <sup>m</sup> = Mann-Whitney U test; <sup>c</sup> = chi-squared test, <sup>f</sup> = fisher's exact test, *patients who pulled out SEEG electrodes before monitoring was complete were excluded<br>IQR: interquartile range; SD: standard deviation; FA: focal aware; FIA: focal impaired awareness; FBTC: focal to bilateral tonic clonic; Max: maximum |  |  |  |  |

### Curative Focal Resective Surgery and Outcome

Of the 12 patients with a single SEEG SOZ, 10 underwent resective surgery with a curative intent (83.33%): 6/8 (75%) TLE patients and 4/4 (100%) E-TLE patients. All surgeries in the TLE group were standard temporal lobectomies. The E-TLE group included a lesionectomy (one patient with a frontal focal cortical dysplasia (FCD) who required SEEG and resective surgery twice), and non-lesional corticectomies. Of the two patients with a single SEEG SOZ who did not go on to have curative surgery, one was lost to follow-up, while the other was an initial TLE patient whose SEEG SOZ was discordant with the MRI brain lesion, leading to a palliative procedure of the lesion instead.

When considering the original sample of patients with BI/U scalp EEG, 6/55 (10.91%) TLE patients and 4/29 (13.79%) E-TLE patients underwent curative surgery. Long-term (> 1 year) Engel outcomes were available for 6/10 (60%) patients: Engel I: one patient (one TLE), Engel II: one patient (one E-TLE), Engel III or IV: four patients (unspecified TLE or E-TLE).

Overall, the estimated probability of undergoing curative resective surgery was 12% and of subsequently achieving an Engel I outcome was 1.2%. (Figure 3)

**Figure 3.**
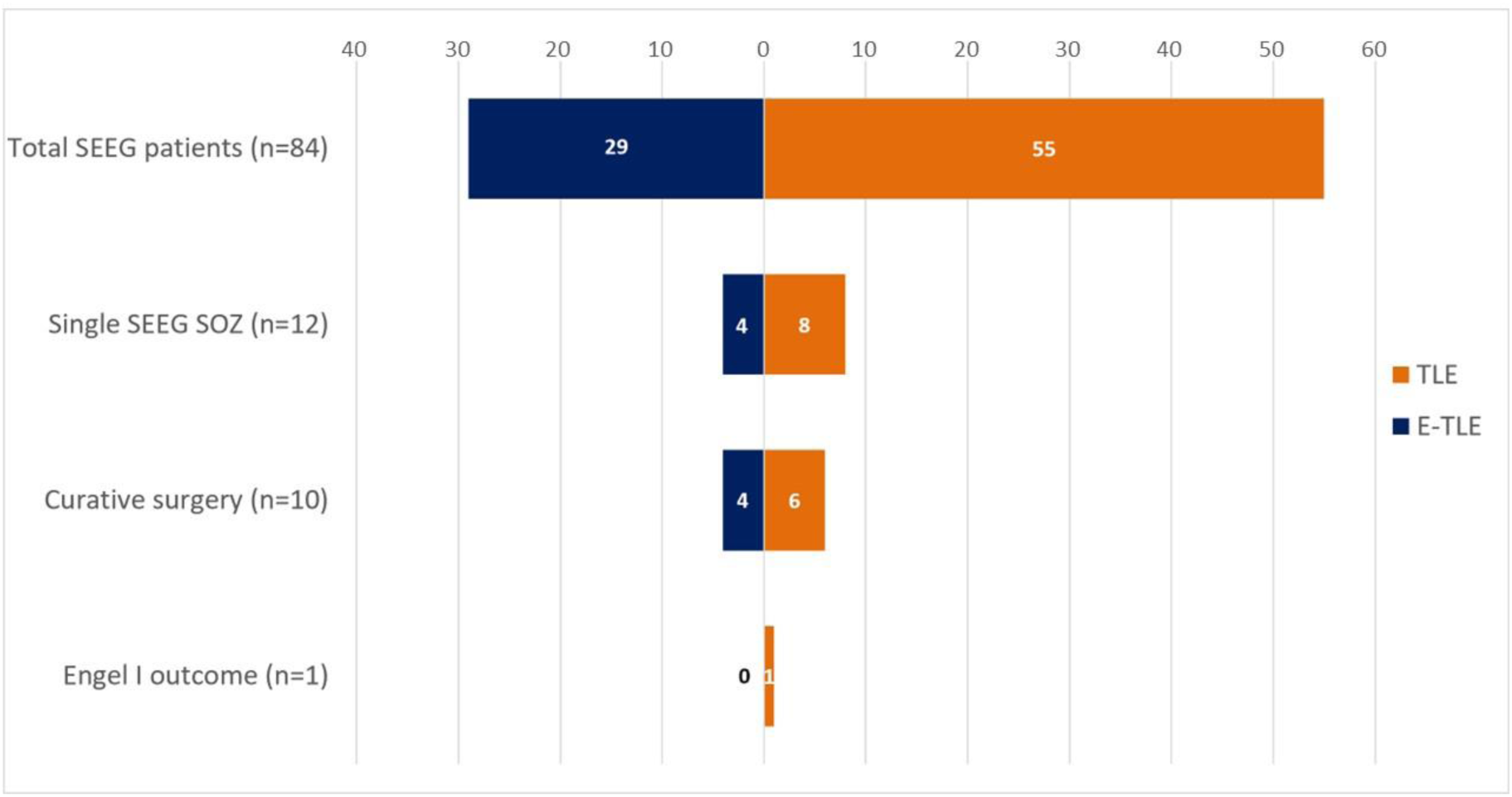
Funnel chart of patients found to have a single SEEG SOZ with subsequent focal surgery and Engel 1 outcome. Numbers have been divided into the TLE and E-TLE group, and demonstrate the low likelihood of finding a single SEEG SOZ in this patient cohort, with even fewer patients going on to have curative surgery and a subsequent Engel 1 outcome. SEEG= stereo-electroencephalography; SOZ= seizure onset zone.

### Palliative Focal Resective Surgery and Neuromodulation

Palliative surgery was offered to 22 patients (one TLE patient with a single SOZ and 21 with multiple SOZs). Of these, 12 were TLE patients (54.54%), half of whom SEEG confirmed bitemporal lobe epilepsy, three fell into the category of TLE-plus while the remaining three had multifocal SEEG SOZs. Long-term Engel outcomes were available for 17/22 (77.27%) patients: Engel I: six patients (three TLE), Engel II: two patients (both TLE), Engel III–IV: nine patients (four TLE). There were no significant outcome differences between TLE and E-TLE groups (p > 0.200). Figure 4 details the Engel outcomes against the duration of follow-up following an intervention for the entire cohort.

**Figure 4.**
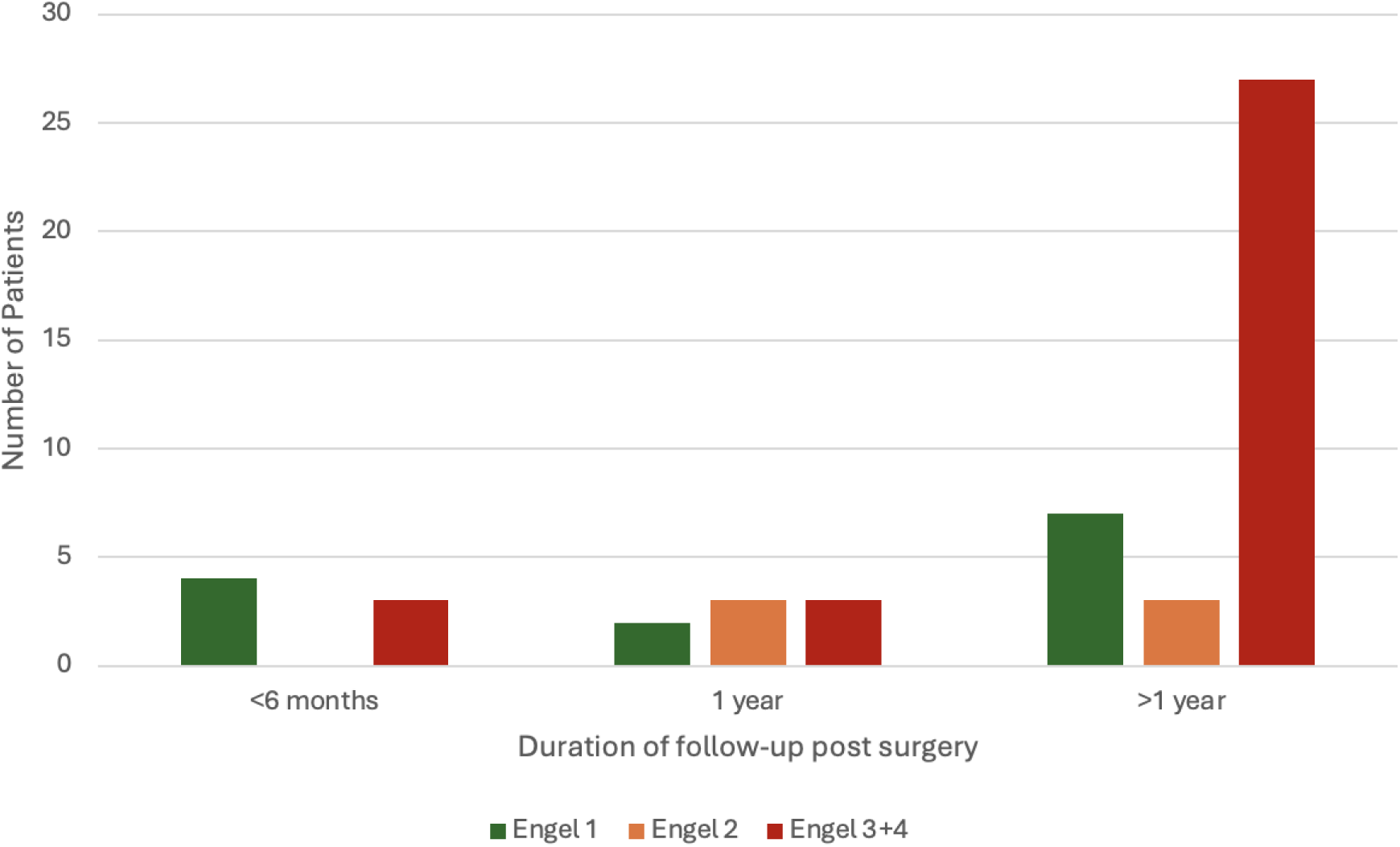
Grouped Bar Chart of the Engel outcomes shown against the duration of follow-up following an intervention for the entire cohort.

Engel I outcomes were achieved in 7% (*n=*6) of these patients. Four of these six patients had concordant MRI brain lesions, with palliative surgeries guided by wide resective margins and awake cortical mapping. A comparison of variables between the palliative and curative surgery groups was limited by small sample sizes.

Neuromodulation included vagus nerve stimulation (VNS), deep brain stimulation and responsive neurostimulation systems (RNS) (details in Table 2). It was performed in 42/84 patients (50%): 28 with TLE and 13 with E-TLE. Among them, 40 had BI scalp EEG SOZs, and two had a single SEEG SOZ. Neuromodulation was offered to 35 patients initially after their SEEG investigations, and a VNS was inserted after failure of focal resective surgery in five patients (two of which were originally performed with curative intent).

### Blinded Validation Sub-study Results

In 72/84 patients (85.71%), SEEG investigations did not identify a resectable target likely to yield seizure freedom. Surgical planning was retrospectively simulated for the 12 patients with a single SOZ. (Supplementary material) Based on phase I data alone, the epilepsy surgery conference recommended proceeding to an SEEG investigation for 6/12 patients, directly to a curative resection in two, neuromodulation for three and a palliative resection in one. The team were then presented with the results of the SEEG implantation, but this did not impact a change in their management/decision in all six cases where SEEG had not originally been recommended. (Figure 5)

**Figure 5.**
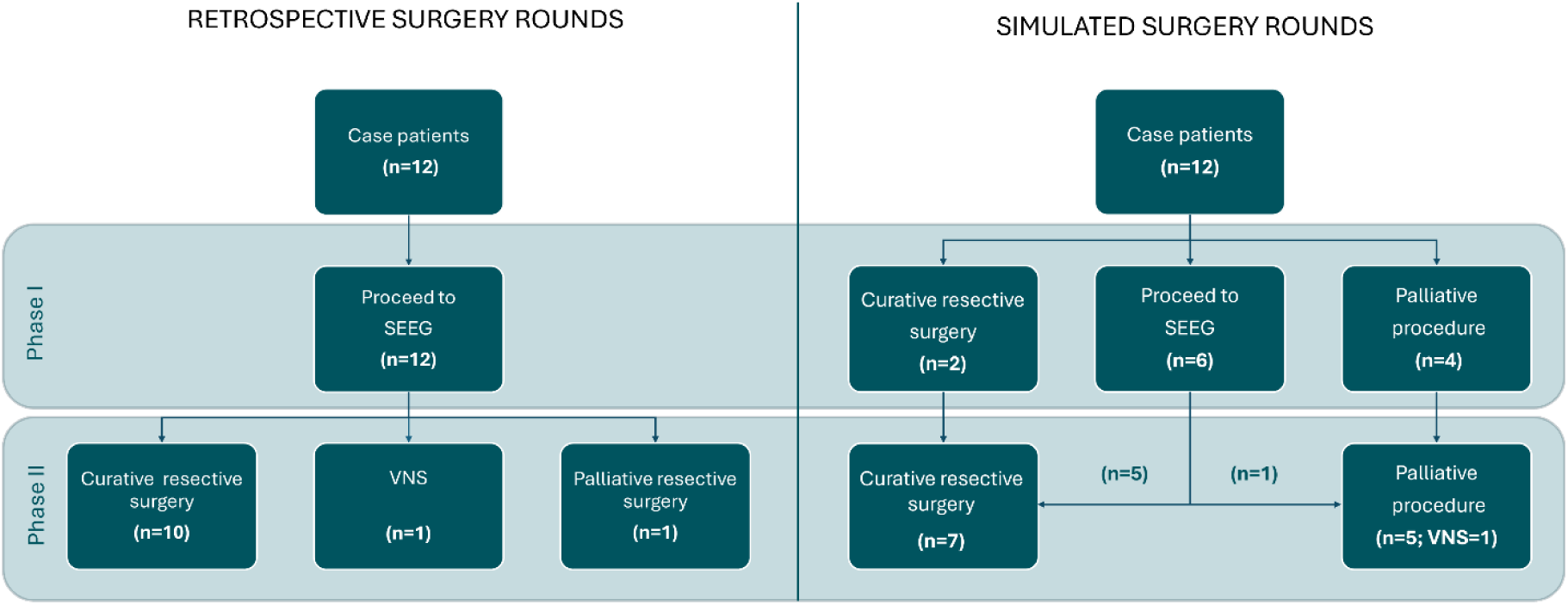
Diagram summarizing the decisions made at the actual epilepsy surgery conference (retrospective surgery rounds) versus the simulated epilepsy surgery conference. The first tier represents the decisions made after being presented with only phase I investigations, while the second tier indicates decisions after phase II investigations were presented. The panel did not change their decisions on any of the SEEG cases once provided with the actual SEEG results, interventions and outcomes. SEEG= stereo-electroencephalography; VNS= vagal nerve stimulation.

In summary, the epilepsy surgery conference determined that in 50% of cases SEEG would not have been pursued, and in 25% patients would have been offered either curative or palliative surgery directly, without proceeding to invasive monitoring.

### Seizure Freedom After SEEG Alone

Additionally, two patients achieved long-term Engel 1 outcomes following an SEEG implantation alone. One was a non-lesional TLE patient who had bitemporal SOZs noted on SEEG, and the other was an E-TLE patient with evidence of bilateral periventricular nodular heterotopia. Despite this, following the SEEG implantation and with an adjustment of one ASM, the patient enjoyed a documented Engel 1 outcome of eight years.

## Discussion

This study, which included 33% of all SEEG cases over 13 years, demonstrates that only 14% of patients with bilateral independent or unclear seizure onset on scalp EEG were found to have a single SOZ after an SEEG investigation. Of these, 83% (less than 15% of the original group) proceeded to curative surgery, yet only one patient achieved a long-term Engel I outcome (1.2%). These findings suggest that BI/U scalp EEG SOZs are associated with a low likelihood of identifying a resectable focus and achieving seizure freedom, and that SEEG with the intent to offer curative surgery may not be warranted in this subgroup. Furthermore, in our blinded decision-validation sub-study, the epilepsy surgery conference would not have proceeded to SEEG in 50% of cases, and in 25% would have directly offered resective surgery. While this result reflects the subjectivity of pre-surgical decision-making, it also underscores the potential to streamline surgical pathways and avoid invasive monitoring in this subgroup.

Scalp EEG remains a central component of the phase I presurgical evaluation, owing to its high temporal resolution and widespread accessibility across epilepsy surgery centers. However, its limitations—most notably low spatial resolution and inability to reliably detect subcortical activity—restrict its utility in precisely localising the epileptogenic zone. ^17^ Composite scoring systems to predict the diagnostic yield of SEEG by integrating multiple clinical variables are being proposed and validated, ^18^ but their application in real-world settings can be challenging due to complexity. In contrast, our findings suggest that a single, easily identifiable variable—BI/U ictal onset on scalp EEG—may offer a pragmatic and clinically relevant tool to guide SEEG candidate selection in a subset of patients who are not uncommon in practice.

Importantly, progression to curative resective surgery following SEEG typically requires the identification of a single, well-defined SOZ that aligns with other clinical and investigational data. ^19^ In our cohort, patients with multifocal or non-localisable SOZs on SEEG uniformly proceeded to palliative procedures or neuromodulatory interventions, consistent with previous studies reporting similar decision-making outcomes. ^11, 19^ Advances in SEEG have broadened surgical possibilities for patients with complex, multifocal epilepsies including with conditions once deemed unsuitable for resection, such as tuberous sclerosis^20^ and polymicrogyria. ^21^ Despite these gains, there has also been a parallel rise in SEEG investigations that ultimately do not lead to resective surgery. ^5^ More recently, there has been growing interest in the potential of SEEG to support alternative therapeutic approaches, such as RF-TC or individualized neuromodulation. ^22–24^ However, these applications fall outside the scope of our present analysis. Importantly, all SEEG implantations in our cohort were undertaken with the primary goal of identifying a single SOZ and achieving seizure freedom.

### Value of SEEG when BI/U Scalp Seizures are Present

While resective surgery may remain appropriate in selected TLE patients with BI/U ictal onset on scalp EEG, our findings indicate that SEEG does not provide additional lateralizing information to meaningfully influence surgical decision-making in this subgroup and represents an unnecessary invasive procedure. Several mechanistic hypotheses have been proposed to justify palliative resections in such cases. These include the concept of secondary epileptogenesis, ^25^ disconnection of propagation pathways, ^26^ and contrasting “mutual facilitation” between the two epileptogenic temporal lobes, in which case removing either may be effective. ^27^ Lastly, “burned-out hippocampus” syndrome has been proposed as an electro-clinical variant which might be responsible for the appearance of bitemporal ictal onset patterns on scalp EEG. ^28^ Notably, in all these scenarios, SEEG is unlikely to offer additional benefit in determining surgical laterality, a notion further supported by the increasing use of RNS in such contexts. ^29^

While there have been a number of studies reporting seizure freedom rates of up to 67% in patients with bitemporal scalp SOZs who went on to have a unilateral temporal resection following invasive EEG studies, ^25, 30, 31^ the degree of seizure lateralization contributed minimally to surgical candidacy in these cohorts. RNS-based chronic ambulatory EEG has helped overcome key limitations of inpatient invasive monitoring, and has demonstrated how monitoring periods of up to eight months are needed before clinical decisions can be made on estimated seizure laterality ratios. ^29^ In our cohort, only 14.55% of TLE patients went on to have a single SOZ found on SEEG during the period of recording. All underwent a temporal lobectomy with curative intent, yet only one achieved an Engel class I outcome; the remainder continued to experience seizures, consistent with bitemporal lobe epilepsy. Further, RNS was an option often considered during the simulated epilepsy surgery conference; however, it is currently an unavailable treatment in Canada.

Several factors may account for BI/U scalp EEG SOZs in E-TLE patients. These include the possible large size of the SOZ, the rapid propagation of the ictal discharge, the presence of focal or generalized attenuation seen with focal-onset seizures and the location and depth of the source generator in, for example, mesial and/or deep sulci. ^32^ There are also network-specific epilepsies originating from certain anatomical areas which demonstrate interhemispheric connectivity, such as perisylvian, tempero-parietal-occipital junction, frontoparietal, and frontal epilepsies, which may demonstrate BI/U scalp EEG SOZs. ^19^ Although these anatomical and physiological features may justify consideration of an SEEG investigation with the intent of localizing a single SOZ, our findings show that only 13.7% of patients in this subgroup ultimately achieved this outcome. It is plausible that this is an overestimation, and that extended recordings or RNS would have revealed multifocal seizure onsets, especially since all of these patients had a shorter duration of recording compared to those with multifocal SEEG SOZs and more importantly, none of these patients achieved a good post-resective surgery outcome. These results underscore the need for cautious interpretation of apparent SOZ localization in E-TLE when BI/U scalp EEG is present.

In our group of patients, SEEG investigations performed in patients who eventually had multifocal/unknown SEEG SOZs were often characterized by a greater number of implanted electrodes and longer recording durations compared to patients with a single SEEG SOZ (p-value 0.01), and what has been previously documented. This reflects the underlying complexity and diagnostic uncertainty in this subgroup. A large systematic review of SEEG in drug-resistant focal epilepsy reported an average of 11.3 electrodes per patient and a mean recording duration of 10.76 days (range: 2–30). ^33^ Another high-volume centre reported an even higher mean electrode count (13.6 ± 2.6, range: 3–22), although most implantations (84%) were unilateral and the number of electrodes did not appear to influence surgical outcomes. ^34^ However, consistent with prior findings from subdural grid studies and select SEEG studies, ^19, 35^ our results reinforce the notion that higher electrode counts may correlate with more diffuse or ambiguous pre-implantation hypotheses, and in turn, with lower rates of resection or seizure freedom.

Finally, in selected cases, palliative resective surgery may remain appropriate despite multifocality. Resection of a functionally dominant focus may disrupt the broader epileptogenic network, ^36, 37^ or only a subset of SOZs may correlate with clinically relevant seizures—as was seen in three patients in our cohort—making them viable targets for palliative intervention. ^38^ Importantly, our data suggest that SEEG is not essential to make this decision. These cases were characterized by visible lesions on MRI in the majority (*n* = 4) and planned resections with wide surgical margins. These findings are congruent with previous studies, which have suggested improved seizure outcomes in lesional epilepsy. ^39^ Due to small numbers, we are unable to directly compare palliative and curative surgery groups or offer definitive recommendations, but our findings highlight that individualized clinical and imaging data may be sufficient for guiding palliative surgical decisions in some BI/U cases without requiring invasive SEEG.

### Impact of Scalp EEG on Good Surgical Outcomes

Multiple studies have shown that a localized ictal onset on scalp EEG in patients with focal epilepsy undergoing SEEG is strongly associated with favourable surgical outcomes. ^17, 40–42^ These are explained by a clearly lateralized scalp EEG SOZ which more effectively guides the placement of depth electrodes, ^40^ a more confined area of epileptogenic activity favouring a more limited and tailored resection and a slower speed of propagation of the seizure. One study showed that patients with rapid seizure spread were not likely to be seizure free in frontal lobe epilepsy, ^42^ and another showed that if the resection included early spread areas, this was associated with a good surgical outcome. ^43^ Our study highlights that patients with BI/U ictal patterns on scalp EEG have both a lower likelihood of identifying a single SOZ on SEEG and reduced chances of achieving seizure freedom after resective surgery. Only seven patients (8%) attained an Engel class I outcome following focal resection, underscoring the clinical relevance of scalp EEG patterns in predicting both the diagnostic utility of SEEG and surgical prognosis. ^44^

The limitations of this study are primarily related to its retrospective, single-centre design, which may limit generalizability. The blinded decision-validation sub-study, while innovative, was conducted with a limited number of reviewers in a simulated environment, which may not fully replicate real-world clinical decision-making. Nonetheless, the study has notable strengths, including a relatively large and well-characterized cohort drawn from a high-volume epilepsy surgery centre. These findings are likely to have clinical relevance for similar tertiary centres. Moreover, the incorporation of a novel blinded decision-validation model to evaluate the added diagnostic value of SEEG in this specific subgroup reinforces our retrospective observations.

In conclusion, patients with bilateral independent or unclear ictal onset on scalp EEG comprised a substantial proportion of SEEG implantations at our center, yet demonstrated a low probability of identifying a single SOZ. Even among those in whom a single SOZ was delineated and who proceeded to focal resective surgery with curative intent, sustained seizure freedom was rare. Although several hypotheses have been proposed to explain why scalp EEG ictal onset may appear BI/U, our findings demonstrate that in reality, this phenomenon corresponds to a well-defined underlying mechanism in only a minority of patients. Given the financial costs, potential surgical morbidity and broader healthcare system burdens associated with SEEG investigations, the presence of a BI/U scalp EEG pattern should prompt critical reassessment of the rationale for invasive monitoring in this subgroup. If SEEG is still considered, it’s essential that patients are counselled about the limited likelihood of both a clear surgical target and a successful outcome. Our findings will help refine how we select candidates for SEEG and avoid unnecessary interventions in a complex, yet large, subgroup.

## Data Availability

All data produced in the present study are available upon reasonable request to the authors

## Acknowledgements

The authors would like to thank Ms Suzan Brown for her assistance with the ethics application of this study, as well as the clinical team for their input. AJS would like to acknowledge the International Federation of Clinical Neurophysiology who awarded her with an Education Award, during the time which she conducted this study.

## Supplementary material

Methods: List of all demographic data and variables collected for included cases.

